# Development of a Novel Dynamic Leak Model to Simulate Leak for Performance Testing of Manual Neonatal Resuscitation Devices. Does Leak Matter? A Bench Study

**DOI:** 10.1101/2024.09.12.24313593

**Authors:** S Morakeas, M Hinder, T Drevhammar, V Gruber, A McEwan, M.B Tracy

## Abstract

**Background:** Newborn resuscitation is commonly performed in the presence of face mask leak. Leak is highly variable, pressure dependent and often unrecognised. The effectiveness of resuscitation devices to deliver adequate inflations in the presence of leak is unknown. Bench models simulating continuous leak have disadvantages of not accurately reflecting leak occurring during clinical resuscitation. A dynamic leak model based on pressure release valves was thus developed.

**Aim:** To assess self-inflating bag (SIB) and T-piece resuscitator (TPR) ventilation performance in the presence of dynamic (DLM) compared to continuous (CLM) leak models in a bench study.

**Method:** Five predefined leak levels were tested for each leak model (0-87%). Resuscitation devices were connected to a test lung (compliance 0.6 mL/cmH_2_O) and respiratory parameters were measured using respiratory function monitors before (patient interface) and after (actual) an induced leak at 40, 60, 80 inflations/min.

**Results:** 3,600 inflations were analysed. DLM showed a decrease in actual tidal volumes from 0%-87% leak with tidal volume differences (SIB 4.8mL, TPR 2.9mL), contrasting to minimal change for CLM (SIB -0.6mL, TPR 0.3mL). CLM demonstrated larger differences between patient interface and actual leak. The absolute difference at 60 inflations/min at 87% leak were SIB 37.5%, TPR 18.2% for CLM compared to SIB 4.6%, TPR 1.4% for DLM.

**Conclusion:** CLM may underestimate the impact of resuscitation device performance with poor correlation between patient interface and actual delivered volume. DLM demonstrates several advantages with more accurate representation of face mask leak and will prove useful in modelling all systems delivering PPV.

- **What is already known on this topic?**
  - Mask leak is common, variable and often unrecognised during newborn resuscitation.
  - Excessive or insufficient tidal volumes may cause newborn lung and brain injury.
  - Current leak models used to test ventilation devices have a poor fidelity in modelling clinical mask leak.
- **What this study adds?**
  - Development of a novel dynamic leak model to enhance performance testing of resuscitation devices by representing pressure dependent leak.
  - Highlights the dynamic nature of leak during clinical resuscitation.
  - Relates RFM displayed leak reliably determining delivered lung volume with leak levels to 86%.
- **How this study might affect research, practice or policy?**
  - The use of dynamic leak model as a higher fidelity simulation tool for the testing of not only resuscitation devices but also ventilators and RFMs.
  - Further research will determine the ability of currently used ventilation devices to perform in the presence of dynamic leak.

## INTRODUCTION

Globally approximately 5-10% of term infants and 20% of preterm infants do not breathe at birth.^1-4^ These infants require resuscitation with positive pressure ventilation (PPV). The delivery of PPV via a facemask using a self-inflating bag (SIB), T-piece resuscitator (TPR) or flow inflating bag is recommended in the 2023 ILCOR guidelines.^2^ The reliability of these devices to deliver consistent adequate tidal volumes and inflation pressures to newborn infants is critical for effective resuscitation.^5, 6^

Performing PPV can be a challenge even for experienced clinicians. Poor technique and face mask leak has been described in a meta-analysis to result in a median mask leak of 29% and may result in substantial fluctuations in delivered ventilation.^7-13^ This can result in inadequate or excessive tidal volumes (Vt) which can cause secondary brain and/or lung injury.^8, 14, 15^ Therefore, it is important to determine how the performance of resuscitation devices used to deliver PPV are impacted by the presence of mask leak. This requires benchmarking with a mechanical model that best simulates face mask leak that occurs during resuscitation at birth.

The effect of leak on a resuscitation device’s ability to deliver volumes and pressures have been evaluated with simulated leak in vitro.^16-19^ Currently published bench models use a continuous leak model (CLM) where leak is present throughout a ventilation cycle.

The CLM was used by Hartung et al. in an in vitro study investigating the performance of resuscitation devices at leak levels (ranging from 0-87%) simulated by different lengths of silicone tubing creating different resistances.^19^ In this model the delivered tidal volume increased with leak.^19^ This result appears counter-intuitive to what is observed clinically and also questions the use of respiratory function monitoring to guide mechanical ventilation. Their findings were explained as an effect of increased driving pressures (Δp = peak inspiratory pressure (PIP) – positive end expiratory pressure (PEEP)) due to more pronounced reductions in PEEP. Other possible factors including the RFM measurement of volume and flow patterns in the leak tubing during deflation were not discussed.

Gomo et al. hypothesised that leak would be dynamic and nonlinear in real-life resuscitations.^20^ Leak would rise and fall directly related to changes in airway pressure and applied mask, shape, size, position, hold and force required.^20^ This was supported in several studies where mask leak has been described as highly variable and pressure dependent.^16, 21-24^

The discrepancy between CLM and real-life experience could potentially be solved with the development of a dynamic leak model (DLM) where leak is controlled by a pressure limiting valve. Leak predominantly occurs at higher pressures and thus is described as pressure dependent leak. This hypothesis led us to the design of a novel dynamic leak model aimed to better simulate leak during PPV with manual resuscitators.

This study aims to compare the performance characteristics of a new DLM to a CLM in a mechanical model simulating newborn resuscitation with a TPR and SIB in the presence of a range of leak levels.

## METHOD AND DEVICES

### Manual Resuscitation Devices and Settings

The performance of two leak models were compared using two manual resuscitation devices: 1. Self-inflating bag (SIB) (Ambu reusable Mark VI, Ambu, Copenhagen, Denmark) with a PEEP valve (Ambu disposable #199 102 001) and 2. T-piece resuscitator (TPR) (Neopuff, Fisher & Paykel Healthcare, Auckland, New Zealand). The set-up is described in Figure 1.

**Figure 1:**
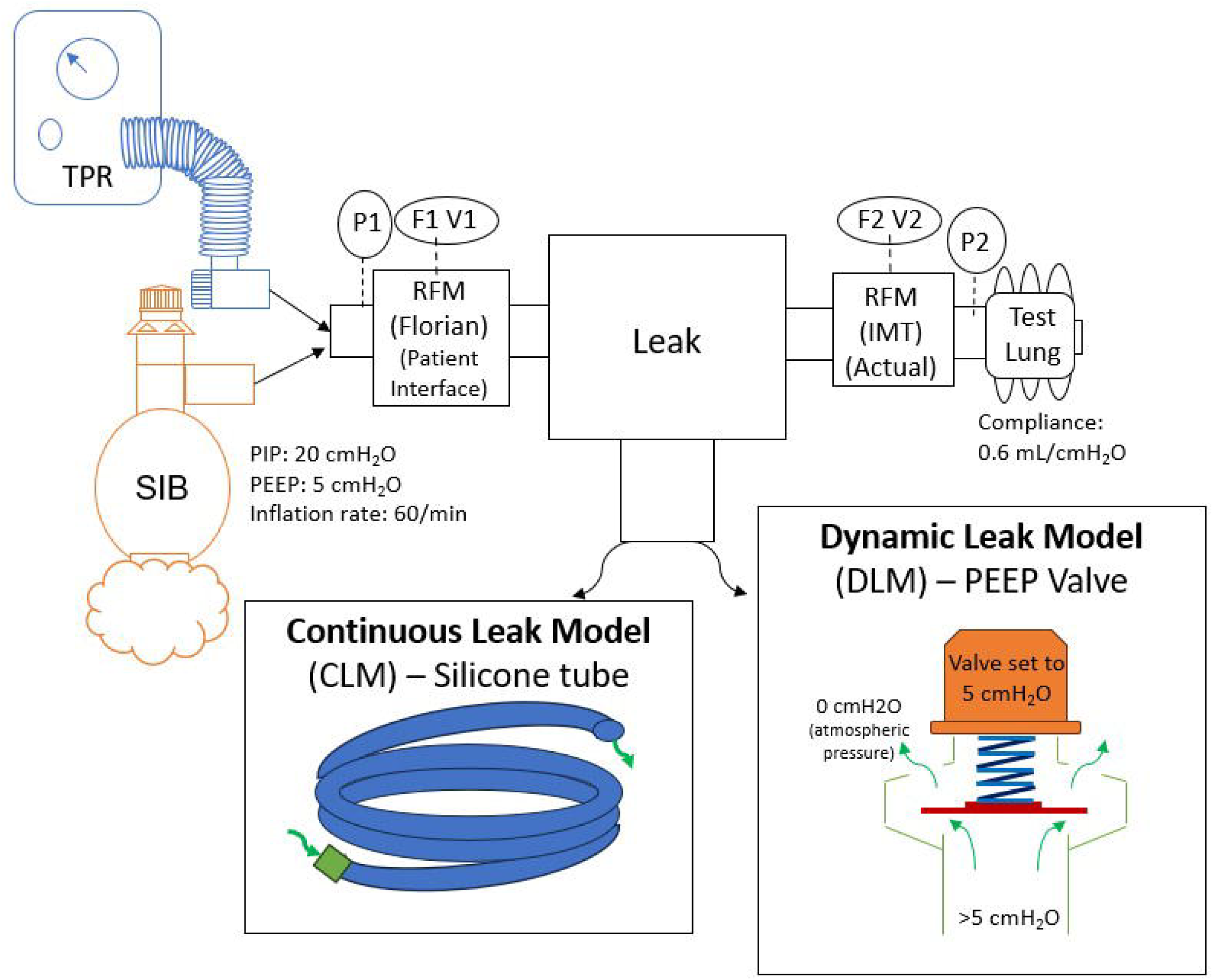
Experimental set-up. The performance of a self-inflating bag (SIB) and t-piece resuscitator (TPR) in the presence of a continuous and dynamic leak models were investigated. The pressure (P), flow (F) and volume (V) were measured at two positions, before an induced leak at the patient interface position and after the induced leak at the test lung (actual) position.

Given the potential fluctuations in hand squeezing the SIB, the SIB device was automatically operated using a robotic mechanism designed to simulate a two-finger bag squeeze previously developed by Tracy et al. ^5^ The T-piece device was manually operated by a single experienced operator (gas flow of 10 L/min), using a metronome to guide the inflation rates. Peak Inspiratory Pressure (PIP) was set at 20 cmH_2_O, and PEEP of 5 cmH_2_O at zero leak across three inflation rates (40, 60 and 80 inflations/min) with an inspiratory and expiratory ratio of 1:1.

### Measurement Devices

A Florian RFM (Acutronic, Medical Systems AG, Zug, Switzerland) was used to measure the ventilation parameters delivered to the system before an induced leak and was referred to as the ‘Patient Interface’ values. A ventilation analyser (FlowAnalyser PF-300 IMT Medical, Buchs, Switzerland) was used to measure parameters delivered to a test lung (compliance of 0.6 ml/cmH_2_O) (Test Lung 191, Maquet, Rastatt, Germany) after an induced leak and was referred to as ‘Actual’ values. The parameters from the patient interface were compared to the actual parameters and used to evaluate the two different leak models.

Before data collection the Florian RFM was calibrated (airway pressure, flow and volume) and measurements verified by a traceable reference ventilation analyser (PF-300 IMT Medical, Buchs, Switzerland) and a precision syringe (Hans Rudolph 5520 10mL). The Florian was within the manufacturer’s stated accuracy specifications. Each leak was then introduced incrementally starting at 0% for each data collection sequence.

### Leak Models

Two leak models were used to simulate CLM and DLM. Five leak levels were predefined at 0%, 22%, 46%, 68% and 87% reproducing the levels used by Hartung et al. reported at 60 inflations/min using their definition of leak.^19^ Leak models tested were:

1. The CLM, using tubing of known lengths and internal diameters.^19, 25^ We used Tygon^®^ tubing with an internal diameter of 1.6mm and outer diameter 4.8 mm cut to lengths presented in Supplementary Table 1.^19, 25^ The tubes were connected using a male luer lock connector (ID: 2 mm) with one end open to the atmosphere. The tube resistance will generate a continuous leakage during inflation and deflation. To simplify data collection several tubes lengths were tuned to match each required leak level for each resuscitation device at an inflation rate of 60 inflations/min.
2. The DLM was developed using PEEP valves (Ambu #199 102 001) set to release at differing pressures (0-20 cmH_2_O) during inflation. The pressure levels were set at a value that produced the same level of leaks. The valve pressure levels were set individually prior to each test at required rate and settings. A new valve was used for each test. The Ambu PEEP valves have large surface area and thus low resistance pressure to produce the DLM. The leaks occurred mostly at higher pressures during inflation. To simplify data collection several identical PEEP valves were used and adjusted to match each inflation rate and resuscitation device.

## Data Collection

Analyses were performed on the flows, volumes and pressure waveforms to determine the PIP, PEEP, tidal volume inspiration (Vti), tidal volume expiration (Vte) at both the patient interface and test lung (actual). 60 inflations were analysed per combination. The data at the patient interface was collected and analysed using Spectra software (Grove Medical, London, England). The actual data measured at the test lung was collected using the IMT FlowAnalyser Flowlab software (IMTMedical, Bushs, Switzerland) and analyses performed using LabChart (AdInstruments, Dunedin, New Zealand). Leak was calculated as the difference between Vti and Vte or Vt_Actual_ as a percentage of Vti (Equation 1 & 2).

### Leak calculations

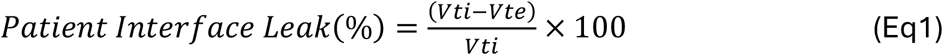

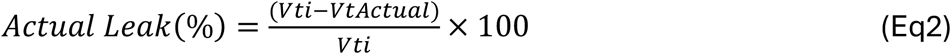

Vti: Tidal volume during inspiration (RFM), Vte: Tidal volume during expiration (RFM), Vt_Actual_: Actual Tidal volume measured at the test lung. The actual leak calculated at the test lung is not affected by expiratory leak and only represents inspiratory leak.

### Statistical Analysis

ANOVA for repeated measures was performed to determine variability and significance of results for all rates and combinations. Summary statistics including mean and standard deviation (SD) was performed to determine variance of pressures, tidal volumes and leak. Negative leak values between -15% and 0% were inspected and re-coded to 0%. Negative leaks less than <-15% were discarded.

## RESULTS

### Overall and Rate

3,600 inflations were analysed. There were small differences between the three inflation rates used. Inflation rates of 60 are presented in detail. Inflation rates of 40 and 80 are available as supplementary data in Supplementary Tables 1, 2 and 3.

### Leak

CLM had larger differences between the leak at the patient interface and actual leak measured at the test lung compared to DLM. The largest absolute difference at 60 inflations/min were recorded with a leak of 87% for CLM: SIB 37.5% and TPR 18.2% compared to SIB: 4.6%, TPR 1.4% for DLM (Table 1, Figure 2). In Figure 2, the relationship between the two leak models, is better correlating for DLM and less so for CLM.

**Table 1:**
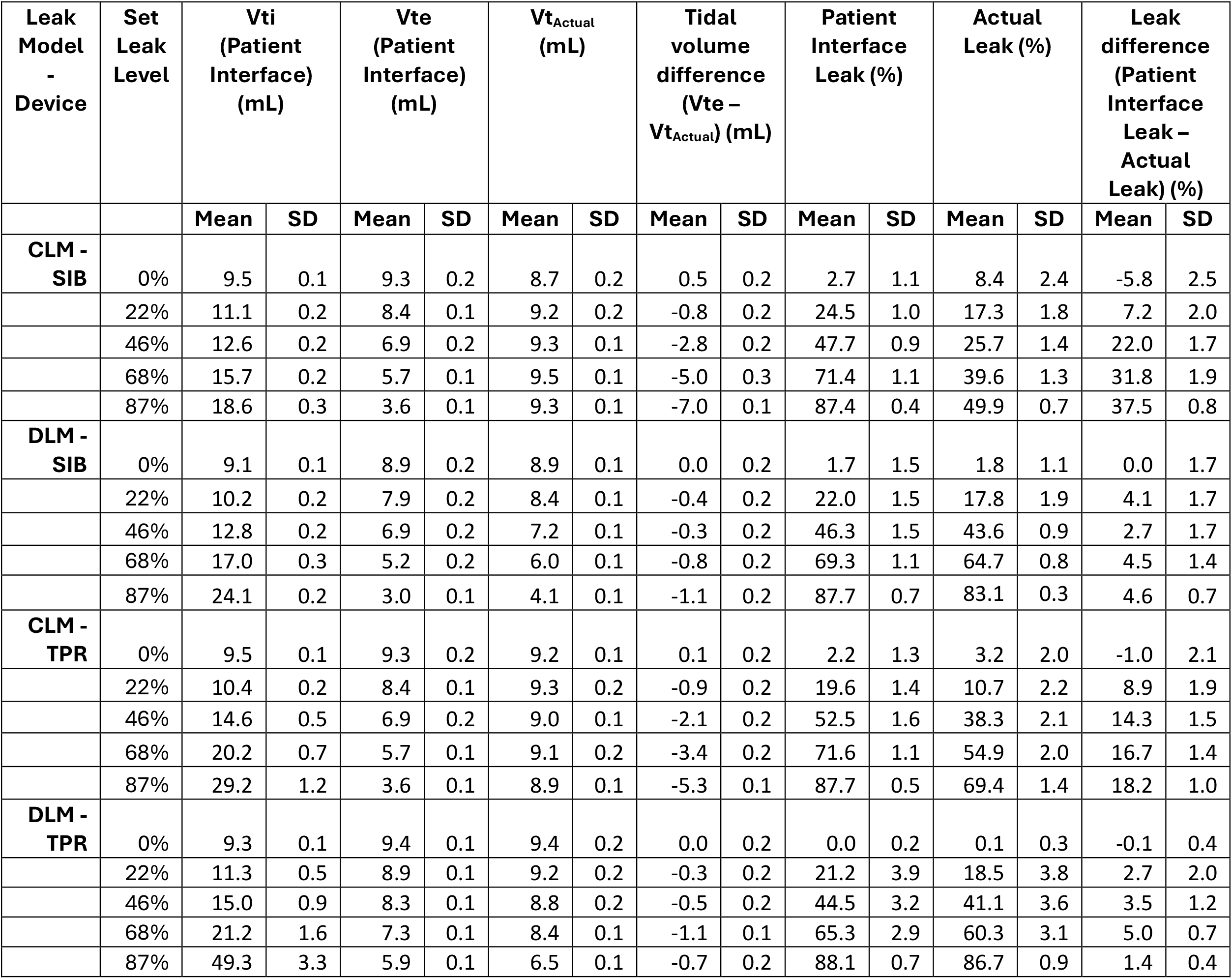
Mean patient interface and actual tidal volumes and leak values for all leak levels at an inflation rate of 60 inflations per minute. Values are presented as the mean and standard deviation (SD).

**Figure 2:**
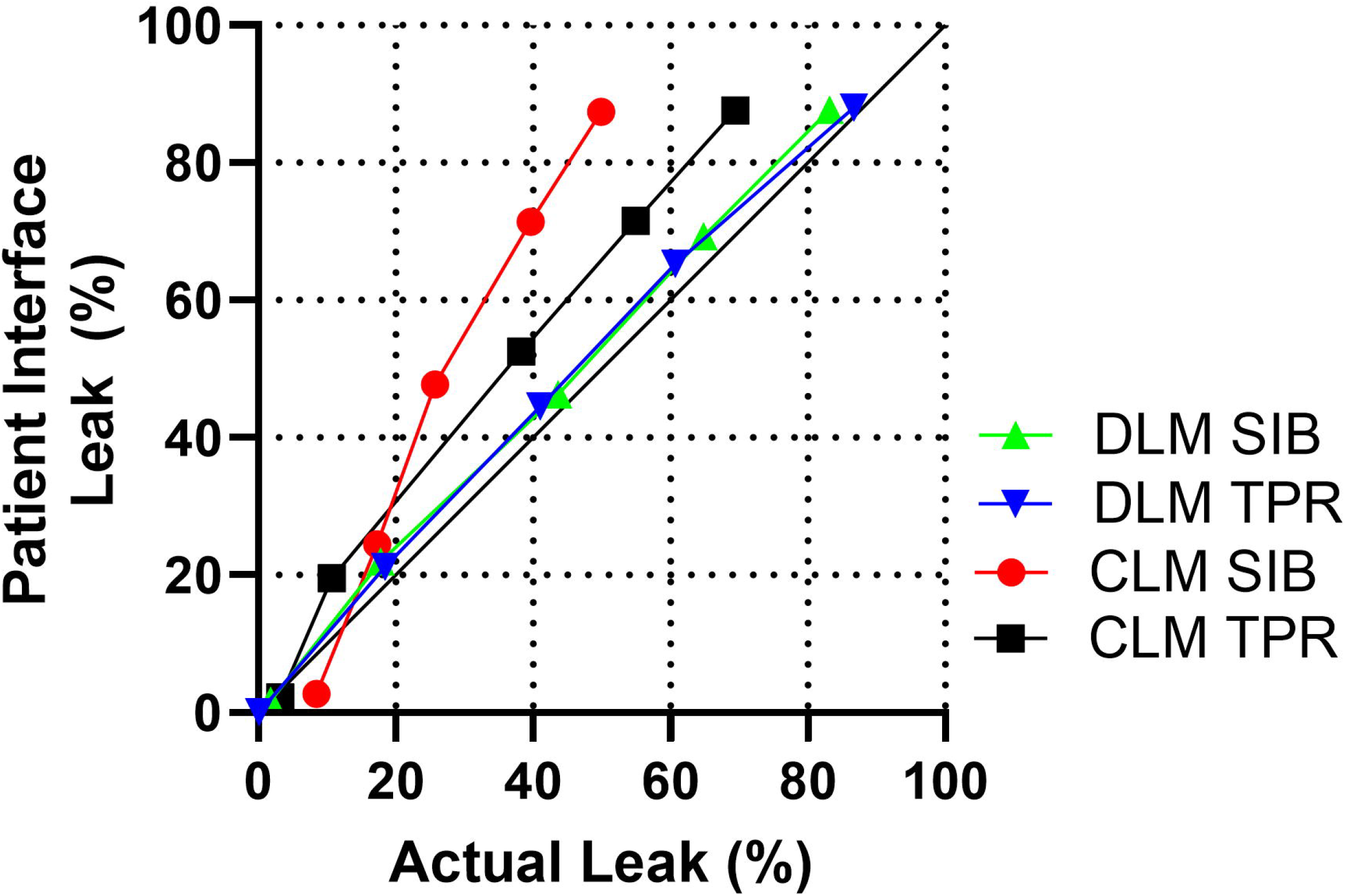
Mean patient interface leak and actual leak for all manual resuscitation devices (Self-inflating bag (SIB) and t-piece resuscitator (TPR)), leak models (continuous leak model (CLM) and dynamic leak model (DLM)) and leak levels for an inflation rate of 60/min.

### Tidal volume

For both devices and leak models, the expired tidal volume at the patient interface decreased as leak increased. The actual tidal volume decreased with increasing leak for the DLM contrasting with tidal volumes remaining relatively constant for the CLM for both devices (Figure 3 A). DLM showed a decrease in actual tidal volumes from 0% to 87% leak (SIB 4.8 mL, TPR 2.9 mL), contrasting to minimal change for CLM (SIB -0.6 mL, TPR 0.3 mL).

**Figure 3:**
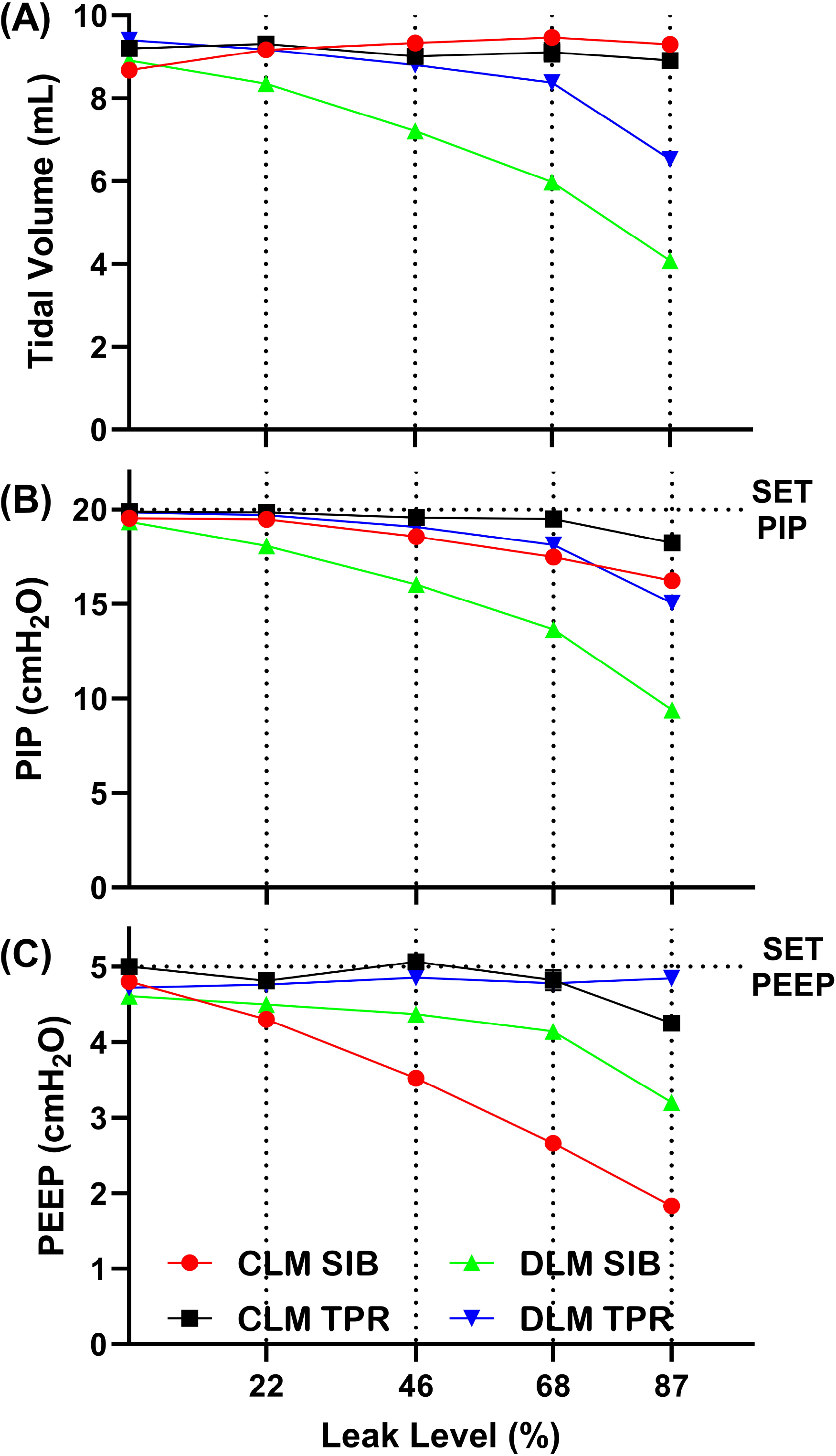
Mean actual respiratory parameters measured at the test lung (A) PIP, (B) PEEP, (C) Tidal volume for all manual resuscitation devices (Self-inflating bag (SIB) and t-piece resuscitator (TPR)), leak models (continuous leak model (CLM) and dynamic leak model (DLM)) and leak levels for an inflation rate of 60/min.

### Pressure

Small differences were observed between the patient interface and actual PIP and PEEP values (Supplementary Table 2). As leak increased the actual PIP and PEEP decreased from the set values for all devices, percentage decrease: DLM (SIB 53.1%, TPR 24.8%); CLM (SIB 18.8%, TPR 8.8%) (Figure 3 B&C, Supplementary Table 2). PEEP remained constant for both leak models when TPR was used. The largest decrease in PEEP was recorded at 87% leak when CLM was used with the SIB with a percentage decrease CLM 63.5% compared to DLM 36.0%.

## DISCUSSION

The variable nature of face mask leak during newborn resuscitation is complex and has previously been simulated using CLM. Compared to this model our novel DLM exhibited a decrease in lung tidal volume as leak increased (Figure 4). This finding is closer to clinical experience with modern ventilators and is consistent with the decrease in tidal volume with increasing leak reported in clinical and in vitro studies.^8, 20, 26-28^ Several studies investigating endotracheal tube and laryngeal mask leak have also reported leak as variable and could be in support of using a dynamic leak model to simulate clinical mask leak.^16, 29-31^

**Figure 4:**
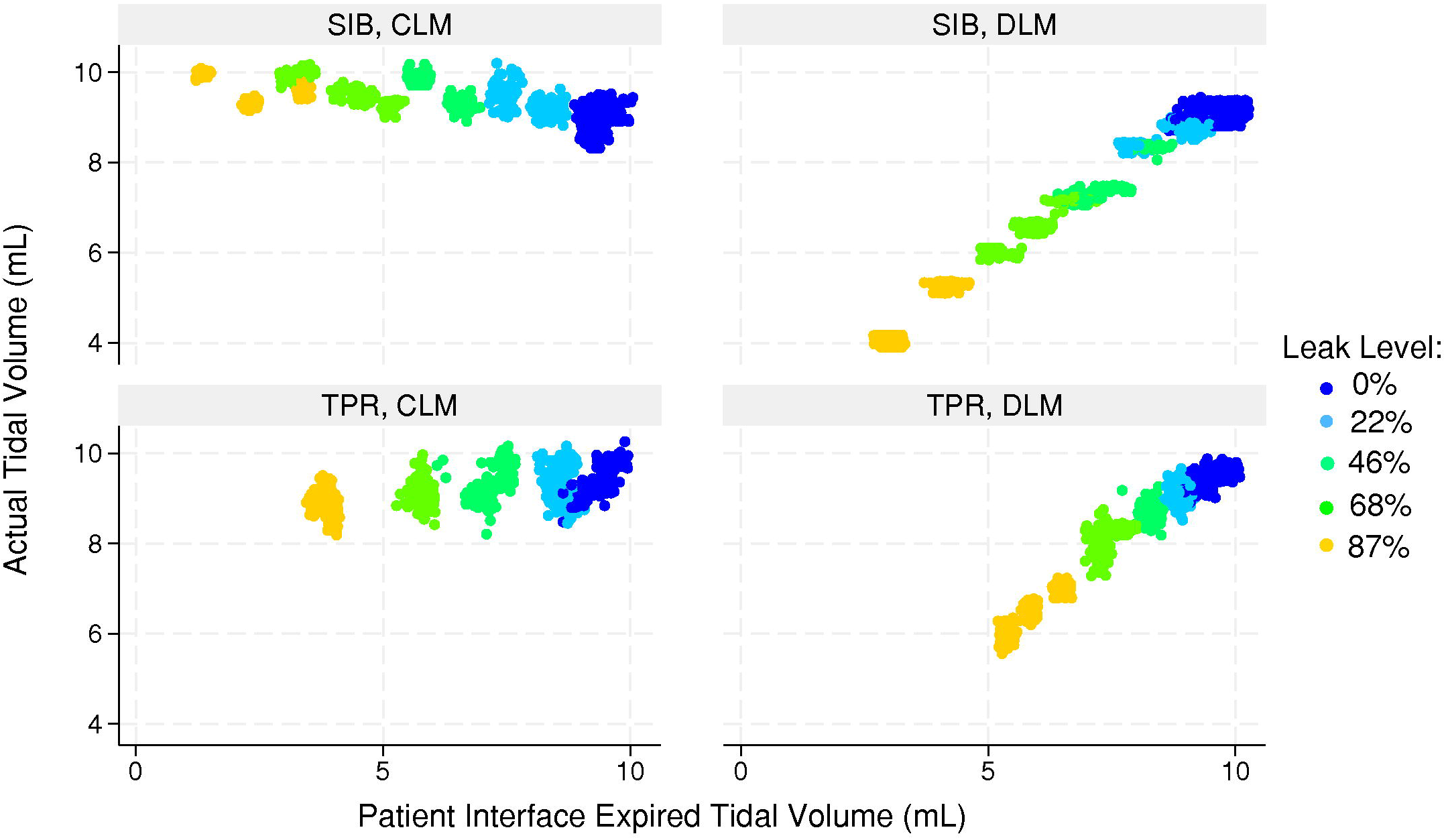
Expiratory tidal volume at the patient interface (Vte) versus the actual tidal volume (Vt_Actual_) for all rates, device and leak model combinations separated into model and device. Different colours represent the five different leak levels (0, 22, 46, 68, 87%).

The pressure dependent nature of leak results in a greater leakage during the inflation phase of the breathing cycle. In our study this was observed for the DLM with small differences between Vt_Actual_ and Vte at the patient interface (Figure 3 A).^19, 22^ This validates the DLM as a pressure dependent leak and indicates that it is a valuable tool to simulate clinical leakage.

The CLM data observed in our study for PIP, PEEP and VT show similar findings to those of Hartung et. al., including the minimal effect on lung volumes with increasing leak.^19^ This finding is interesting and not experienced clinically.^20^ The observed lack of reduction in volume despite leak was explained by Hartung et.al as related to unchanged driving pressure (Δp) with increased leak since both PIP and PEEP pressures were reduced.^19^ This behaviour was observed for CLM in our study, consistent with Hartung’s study. We do not consider Δp with CLM as the entire explanation for unchanged lung volumes with system leaks as high as 87%. ANOVA adjusted means for Δp showed reduced pressure from 15.1 cmH_2_O with 0% leak to 11.4 cmH_2_O at 86% leak adjusting for model, device and rate. The R^2^ coefficient of 0.65 indicated other factors may be important and not in the model such as deflation cycle entrainment of volume. Mechanical ventilators use ventilator specific leak compensation algorithm to correct tidal volume measurements and adjust delivered flows and pressures to achieve required tidal volumes with the best performing ventilators having an accuracy of ±10% at high leak values.^32^ This is not possible in resuscitation using manual devices as these devices rely on the skills of the clinician, face mask technique and crude tidal volume estimations based on chest wall rise. ^6, 7, 33-36^ Therefore, it is important to determine the ability of manual resuscitation devices to perform in the presence of leak.

In our study, PPV provided with TPR was less sensitive to the presence of leak compared to SIB. This confirms the findings by O’Donnell et al. and Hartung et al.^19, 22^ The TPR was able to deliver adequate volumes (6-8 ml/kg) at the largest leak level of 87% in both leak models, with little decrease in set pressures (Figure 3 B&C, Supplementary Table 2).^10, 34^ The continuous flow of the TPR compensates for leak and this same mechanism has been reported to create a greater leak occurrence with SIB.^22^ The ability of an SIB to compensate for leak depends on the size of air reservoir and squeeze distance. SIB was only able to deliver adequate tidal volumes and pressures up to 68% leak. Competent airway management with low leak is desired in newborn resuscitation.

The presented study has several limitations. Bench studies and findings may not be directly applicable to clinical practise. It is possible that combinations of dynamic and continuous mask leak are present during resuscitation. The pressure valves used for DLM were limited to a maximum pressure level of 20cmH_2_O and have been shown to be variable if not adjusted correctly.^37, 38^ Inspiratory and expiratory time constants can have an impact on leak and delivered pressures. Development of complex and controlled leak models such as electronically operated valves are required to further investigate the impact of time constants and combination leaks on system leakage.

The presence of leak and unsatisfactory airway management can result in resuscitation failure. RFMs can assist clinicians in identifying and quantifying leak during a resuscitation at the patient interface. In infants it is impossible to measure respiratory parameters at the lung. Clinicians should be aware that a large leak can create errors in tidal volumes measured at the patient interface and underestimate the actual tidal volume entering the lungs. This is why in the presence of a leak tidal volume is best reported as expired tidal volume as most of the leak occurs during inflation when the PIP is higher. Further investigation is required to quantify the accuracy of RFMs in the presence of variable leak.

A recent systematic review and ILCOR guideline identified that there is no clear definition what size of leak (percentage) during resuscitation is considered clinically significant, with various studies reporting different levels of significance.^10^ A defined level of clinically significant leak will be difficult to determine as leaks are rarely constant. The interpretation and compensation of leak during resucitation is determined by both the clinican’s ability and device characteristics. We believe that developing new strategies to accurately simulate leak is important for improving resuscitation training, device performance and clinical care.

## Conclusions

We have demonstrated that this new dynamic leak model provides more consistent tidal volume and leak information between delivered volumes and volumes measured at the patient interface. This new DLM will be useful for validating manual resuscitation devices and possibly more complex modes in mechanical ventilators such as volume targeting.

## Data Availability

All data produced in the present work are contained in the manuscript and supplementary material.

## Contributions

SM design of study protocol, carried out data collection, data analysis/statistics, drafted the initial manuscript, reviewed, and approved final manuscript.

MBT conceptualised and designed study protocol, performed data analysis/statistics, contributed and reviewed manuscript, and approved final manuscript.

MH conceptualised and designed study protocol, provided theoretical and technical support, contributed and reviewed manuscript, and approved final manuscript.

TD provided theoretical and technical support, contributed and reviewed manuscript, and approved final manuscript.

VG assisted with data collection, review of manuscript, and approved final manuscript. AM provided technical support and reviewed and approved final manuscript.

## Funding

We acknowledge funding support from a PhD Research Grant awarded by Cerebral Palsy Alliance. This research is also supported by an Australian Government Research Training Program (RTP) Scholarship.

## Acknowledgements

We thank Professor Colin Morley for helpful comments that have improved this paper.

## Competing interests

None declared.

## Ethics approval

Ethics approval was not required as this study is a bench study and did not involve human participants.

## Notes

### Competing Interest Statement

The authors have declared no competing interest.

