## Supplementary material for "Development of a Novel Dynamic Leak Model to Simulate Leak for Performance Testing of Manual Neonatal Resuscitation Devices. Does Leak Matter? A Bench Study"

**Supplementary Table 1:** Validation of leak level set for each model, mean and standard deviation (SD) presented for leak values calculated at the patient interface.

| Leak Level | Tube Length (cm) | Tube Resistance ($cmH_2O/L/s$) | Leak (%) at inflation rates: Mean ±SD | | |
|---|---|---|---|---|---|
| | | | **40/min** | **60/min** | **80/min** |
| CLM - SIB | | | | | |
| 0% | - | - | 3.3 ± 1.4 | 2.7 ± 1.1 | 2.0 ± 1.6 |
| 22% | 266.8 | 4495.2 | 39.5 ± 0.9 | 24.5 ± 1.0 | 20.0 ± 1.6 |
| 46% | 85.0 | 1489.2 | 61.6 ± 0.7 | 47.7 ± 0.9 | 43.2 ± 1.2 |
| 68% | 38.5 | 696.0 | 82.4 ± 0.9 | 71.4 ± 1.1 | 61.81 ± 0.5 |
| 87% | 20.0 | 363.6 | 94.1 ± 0.3 | 87.4 ± 0.4 | 79.5 ± 0.4 |
| CLM - TPR | | | | | |
| 0% | - | - | 1.5 ± 1.1 | 2.2 ± 1.3 | 2.0 ± 1.5 |
| 22% | 405.0 | 6999.6 | 25.8 ± 1.7 | 19.6 ± 1.4 | 15.1 ± 1.7 |
| 46% | 108.0 | 2028.0 | 51.0 ± 2.8 | 52.5 ± 1.6 | 44.3 ± 1.2 |
| 68% | 45.0 | 867.6 | 74.4 ± 0.8 | 71.6 ± 1.1 | 68.0 ± 1.1 |
| 87% | 16.7 | 332.4 | 89.1 ± 0.3 | 87.7 ± 0.5 | 83.1 ± 0.6 |
| DLM - SIB | | | | | |
| 0% | | | 0.1 ± 0.3 | 1.7 ± 1.5 | 0.7 ± 1.5 |
| 22% | | | 24.5 ± 1.5 | 22.0 ± 1.5 | 21.9 ± 1.6 |
| 46% | | | 45.0 ± 1.1 | 46.3 ± 1.5 | 44.9 ± 1.3 |
| 68% | | | 68.1 ± 1.0 | 69.3 ± 1.1 | 66.5 ± 1.1 |
| 87% | | | 87.3 ±0.6 | 87.7 ± 0.7 | 86.8 ± 0.8 |
| DLM - TPR | | | | | |
| 0% | | | 2.0 ± 2.5 | 0.1 ± 0.4 | 0.3 ± 0.7 |
| 22% | | | 22.7 ± 2.9 | 21.2 ± 3.9 | 21.6 ± 2.7 |
| 46% | | | 46.1 ± 3.5 | 44.5 ± 3.2 | 47.4 ± 2.7 |
| 68% | | | 67.6 ± 1.9 | 65.3 ± 2.9 | 70.4 ± 20.7 |
| 87% | | | 86.6 ± 2.1 | 88.1 ± 0.7 | 87.9 ± 1.5 |

**Supplementary Table 2:** Actual pressure measured at 40, 60 and 80 inflations per minute. Values presented as the mean and standard deviation (SD) as a percentage.

| Leak Model - Device | Rate (inflations /min) | Set Leak Level | PIP ($cmH_2O$) | | PIP drop from set PIP of 20$cmH_2O$ | | PEEP ($cmH_2O$) | | PEEP drop from set PEEP of 5$cmH_2O$ | | Delta P (PIP – PEEP) | |
|---|---|---|---|---|---|---|---|---|---|---|---|---|
| | | | Mean | SD | Mean | SD | Mean | SD | Mean | SD | Mean | SD |
| CLM - SIB | 40 | 0% | 19.8 | 0.0 | 0.2 | 0.0 | 4.6 | 0.0 | 0.4 | 0.0 | 15.2 | 0.0 |
| | | 22% | 19.2 | 0.0 | 0.8 | 0.0 | 3.8 | 0.0 | 1.2 | 0.0 | 15.4 | 0.0 |
| | | 46% | 18.1 | 0.0 | 1.9 | 0.0 | 2.6 | 0.0 | 2.4 | 0.0 | 15.5 | 0.0 |
| | | 68% | 16.9 | 0.0 | 3.1 | 0.0 | 1.7 | 0.0 | 3.3 | 0.0 | 15.2 | 0.0 |
| | | 87% | 15.7 | 0.0 | 4.3 | 0.0 | 0.9 | 0.0 | 4.1 | 0.0 | 14.7 | 0.0 |
| | 60 | 0% | 19.5 | 0.0 | 0.5 | 0.0 | 4.8 | 0.1 | 0.2 | 0.1 | 14.7 | 0.1 |
| | | 22% | 19.5 | 0.0 | 0.5 | 0.0 | 4.3 | 0.0 | 0.7 | 0.0 | 15.2 | 0.0 |
| | | 46% | 18.6 | 0.0 | 1.4 | 0.0 | 3.5 | 0.0 | 1.5 | 0.0 | 15.1 | 0.1 |
| | | 68% | 17.5 | 0.1 | 2.5 | 0.1 | 2.7 | 0.0 | 2.3 | 0.0 | 14.8 | 0.1 |
| | | 87% | 16.2 | 0.0 | 3.8 | 0.0 | 1.8 | 0.0 | 3.2 | 0.0 | 14.4 | 0.1 |
| | 80 | 0% | 20.0 | 0.0 | 0.0 | 0.0 | 4.7 | 0.0 | 0.3 | 0.0 | 15.3 | 0.1 |
| | | 22% | 19.7 | 0.0 | 0.3 | 0.0 | 4.5 | 0.0 | 0.5 | 0.0 | 15.2 | 0.0 |
| | | 46% | 19.0 | 0.0 | 1.0 | 0.0 | 3.9 | 0.0 | 1.1 | 0.0 | 15.2 | 0.0 |
| | | 68% | 18.0 | 0.0 | 2.0 | 0.0 | 3.2 | 0.0 | 1.8 | 0.0 | 14.8 | 0.0 |
| | | 87% | 17.3 | 0.0 | 2.7 | 0.0 | 2.4 | 0.0 | 2.6 | 0.0 | 14.8 | 0.1 |
| DLM - SIB | 40 | 0% | 19.6 | 0.0 | 0.4 | 0.0 | 4.4 | 0.0 | 0.6 | 0.0 | 15.1 | 0.0 |
| | | 22% | 19.1 | 0.0 | 0.9 | 0.0 | 4.4 | 0.0 | 0.6 | 0.0 | 14.8 | 0.0 |
| | | 46% | 18.3 | 0.0 | 1.7 | 0.0 | 4.4 | 0.0 | 0.6 | 0.0 | 13.9 | 0.0 |
| | | 68% | 15.6 | 0.0 | 4.4 | 0.0 | 4.2 | 0.0 | 0.8 | 0.0 | 11.5 | 0.0 |
| | | 87% | 11.1 | 0.0 | 8.9 | 0.0 | 4.4 | 0.0 | 1.9 | 0.0 | 8.0 | 0.0 |
| | 60 | 0% | 19.3 | 0.1 | 0.7 | 0.1 | 4.6 | 0.0 | 0.4 | 0.0 | 14.7 | 0.1 |
| | | 22% | 18.1 | 0.0 | 1.9 | 0.0 | 4.5 | 0.0 | 0.5 | 0.0 | 13.6 | 0.1 |
| | | 46% | 16.0 | 0.0 | 4.0 | 0.0 | 4.4 | 0.0 | 0.6 | 0.0 | 11.7 | 0.0 |
| | | 68% | 13.6 | 0.0 | 6.4 | 0.0 | 4.1 | 0.0 | 0.9 | 0.0 | 9.5 | 0.1 |
| | | 87% | 9.4 | 0.0 | 10.6 | 0.0 | 4.6 | 0.0 | 1.8 | 0.0 | 6.2 | 0.0 |

| | | | | | | | | | | | | |
|---|---|---|---|---|---|---|---|---|---|---|---|---|
| | 80 | 0% | 19.8 | 0.0 | 0.2 | 0.0 | 4.7 | 0.0 | 0.3 | 0.0 | 15.1 | 0.0 |
| | | 22% | 19.2 | 0.0 | 0.8 | 0.0 | 4.6 | 0.0 | 0.4 | 0.0 | 14.6 | 0.0 |
| | | 46% | 16.9 | 0.1 | 3.1 | 0.1 | 4.5 | 0.0 | 0.5 | 0.0 | 12.4 | 0.1 |
| | | 68% | 15.2 | 0.0 | 4.8 | 0.0 | 4.5 | 0.0 | 0.5 | 0.0 | 10.7 | 0.0 |
| | | 87% | 10.3 | 0.0 | 9.7 | 0.0 | 4.7 | 0.0 | 1.1 | 0.0 | 6.4 | 0.0 |
| CLM - TPR | 40 | 0% | 20.3 | 0.0 | -0.3 | 0.0 | 4.6 | 0.1 | 0.4 | 0.1 | 15.6 | 0.1 |
| | | 22% | 20.0 | 0.1 | 0.0 | 0.1 | 4.6 | 0.1 | 0.4 | 0.1 | 15.3 | 0.1 |
| | | 46% | 19.8 | 0.1 | 0.2 | 0.1 | 4.6 | 0.1 | 0.4 | 0.1 | 15.1 | 0.1 |
| | | 68% | 18.9 | 0.0 | 1.1 | 0.0 | 4.6 | 0.2 | 0.4 | 0.2 | 14.3 | 0.1 |
| | | 87% | 18.1 | 0.0 | 1.9 | 0.0 | 4.1 | 0.0 | 0.9 | 0.0 | 14 | 0.1 |
| | 60 | 0% | 19.9 | 0.0 | 0.1 | 0.0 | 5.0 | 0.1 | 0.0 | 0.1 | 14.9 | 0.1 |
| | | 22% | 19.8 | 0.0 | 0.2 | 0.0 | 4.8 | 0.1 | 0.2 | 0.1 | 15.0 | 0.1 |
| | | 46% | 19.6 | 0.0 | 0.4 | 0.0 | 5.1 | 0.1 | -0.1 | 0.1 | 14.5 | 0.1 |
| | | 68% | 19.5 | 0.0 | 0.5 | 0.0 | 4.8 | 0.1 | 0.2 | 0.1 | 14.7 | 0.1 |
| | | 87% | 18.2 | 0.0 | 1.8 | 0.0 | 4.3 | 0.1 | 0.7 | 0.1 | 14.0 | 0.1 |
| | 80 | 0% | 19.9 | 0.0 | 0.1 | 0.0 | 5.0 | 0.1 | 0.0 | 0.1 | 14.9 | 0.1 |
| | | 22% | 19.7 | 0.0 | 0.3 | 0.0 | 5.1 | 0.1 | -0.1 | 0.1 | 14.7 | 0.1 |
| | | 46% | 19.3 | 0.0 | 0.7 | 0.0 | 4.7 | 0.1 | 0.3 | 0.1 | 14.6 | 0.1 |
| | | 68% | 19.3 | 0.0 | 0.7 | 0.0 | 4.7 | 0.1 | 0.3 | 0.1 | 14.6 | 0.1 |
| | | 87% | 18.2 | 0.0 | 1.8 | 0.0 | 4.3 | 0.1 | 0.7 | 0.1 | 13.8 | 0.1 |
| DLM - TPR | 40 | 0% | 19.9 | 0.0 | 0.1 | 0.0 | 4.7 | 0.1 | 0.3 | 0.1 | 15.2 | 0.1 |
| | | 22% | 19.7 | 0.0 | 0.3 | 0.0 | 5.2 | 0.0 | -0.2 | 0.0 | 14.5 | 0.0 |
| | | 46% | 19.4 | 0.0 | 0.6 | 0.0 | 5.3 | 0.0 | -0.3 | 0.0 | 14.1 | 0.0 |
| | | 68% | 19.0 | 0.0 | 1.0 | 0.0 | 5.3 | 0.0 | -0.3 | 0.0 | 13.6 | 0.1 |
| | | 87% | 17.2 | 0.1 | 2.8 | 0.1 | 5.6 | 0.1 | -0.6 | 0.1 | 11.6 | 0.1 |
| | 60 | 0% | 19.9 | 0.0 | 0.1 | 0.0 | 4.7 | 0.1 | 0.3 | 0.1 | 15.1 | 0.1 |
| | | 22% | 19.7 | 0.0 | 0.3 | 0.0 | 4.8 | 0.1 | 0.2 | 0.1 | 14.9 | 0.1 |
| | | 46% | 19.1 | 0.0 | 0.9 | 0.0 | 4.8 | 0.1 | 0.2 | 0.1 | 14.2 | 0.1 |
| | | 68% | 18.1 | 0.1 | 1.9 | 0.1 | 4.8 | 0.0 | 0.2 | 0.0 | 13.3 | 0.1 |
| | | 87% | 15.0 | 0.0 | 5.0 | 0.0 | 4.8 | 0.0 | 0.2 | 0.0 | 10.2 | 0.0 |
| | 80 | 0% | 20.0 | 0.0 | 0.0 | 0.0 | 4.7 | 0.1 | 0.3 | 0.1 | 15.3 | 0.1 |
| | | 22% | 19.7 | 0.0 | 0.3 | 0.0 | 4.8 | 0.1 | 0.2 | 0.1 | 14.9 | 0.1 |
| | | 46% | 18.7 | 0.0 | 1.3 | 0.0 | 4.8 | 0.1 | 0.2 | 0.1 | 13.9 | 0.1 |
| | | 68% | 17.4 | 0.0 | 2.6 | 0.0 | 4.9 | 0.1 | 0.1 | 0.1 | 12.5 | 0.1 |
| | | 87% | 14.2 | 0.0 | 5.8 | 0.0 | 4.8 | 0.1 | 0.2 | 0.1 | 9.4 | 0.1 |

**Supplementary Table 3:** Mean patient interface and actual tidal volumes and leak values for all leak levels at an inflation rate of 60 inflations per minute. Values are presented as the mean and standard deviation (SD) as a percentage.

| Leak model - Device | Rate (inflations /min) | Set Leak Level | Vti Patient Interface (mL) | | Vte Patient Interface (mL) | | $Vt_{lung}$ Test Lung (mL) | | Tidal volume difference (Vte – $Vt_{lung}$) (mL) | | Patient Interface leak (%) | | Test Lung Leak (%) | | Leak difference (Patient Interface Leak – Test Lung Leak) (%) | |
|---|---|---|---|---|---|---|---|---|---|---|---|---|---|---|---|---|
| | | | Mean | SD | Mean | SD | Mean | SD | Mean | SD | Mean | SD | Mean | SD | Mean | SD |
| CLM - SIB | 40 | 0% | 9.9 | 0.2 | 9.6 | 0.2 | 9.2 | 0.2 | 0.4 | 0.3 | 3.3 | 1.4 | 7.4 | 2.6 | -4.2 | 3.1 |
| | | 22% | 12.3 | 0.2 | 8.6 | 0.2 | 9.6 | 0.3 | -2.1 | 0.3 | 39.5 | 0.9 | 22.1 | 2.4 | 17.4 | 2.6 |
| | | 46% | 14.9 | 0.2 | 7.4 | 0.4 | 9.9 | 0.1 | -4.2 | 0.2 | 61.6 | 0.7 | 33.7 | 1.1 | 27.9 | 1.3 |
| | | 68% | 18.4 | 0.3 | 5.7 | 0.1 | 9.9 | 0.1 | -6.7 | 0.2 | 82.4 | 0.9 | 46.1 | 0.9 | 36.4 | 1.4 |
| | | 87% | 22.6 | 0.3 | 3.8 | 0.1 | 10.0 | 0.1 | -8.6 | 0.1 | 94.1 | 0.3 | 55.8 | 0.5 | 38.2 | 0.7 |
| | 60 | 0% | 9.5 | 0.1 | 9.3 | 0.2 | 8.7 | 0.2 | 0.5 | 0.2 | 2.7 | 1.1 | 8.4 | 2.4 | -5.8 | 2.5 |
| | | 22% | 11.1 | 0.2 | 8.4 | 0.1 | 9.2 | 0.2 | -0.8 | 0.2 | 24.5 | 1.0 | 17.3 | 1.8 | 7.2 | 2.0 |
| | | 46% | 12.6 | 0.2 | 6.9 | 0.2 | 9.3 | 0.1 | -2.8 | 0.2 | 47.7 | 0.9 | 25.7 | 1.4 | 22.0 | 1.7 |
| | | 68% | 15.7 | 0.2 | 5.7 | 0.1 | 9.5 | 0.1 | -5.0 | 0.3 | 71.4 | 1.1 | 39.6 | 1.3 | 31.8 | 1.9 |
| | | 87% | 18.6 | 0.3 | 3.6 | 0.1 | 9.3 | 0.1 | -7.0 | 0.1 | 87.4 | 0.4 | 49.9 | 0.7 | 37.5 | 0.8 |
| | 80 | 0% | 9.4 | 0.1 | 8.9 | 0.1 | 9.2 | 0.1 | 0.1 | 0.2 | 2.0 | 1.6 | 2.8 | 1.8 | -0.8 | 2.2 |
| | | 22% | 10.3 | 0.1 | 8.7 | 0.2 | 9.2 | 0.2 | -0.9 | 0.2 | 19.3 | 1.2 | 10.6 | 1.8 | 8.7 | 2.4 |
| | | 46% | 11.7 | 0.1 | 7.1 | 0.1 | 9.3 | 0.1 | -2.6 | 0.1 | 43.2 | 0.7 | 20.6 | 1.1 | 22.6 | 1.3 |
| | | 68% | 13.5 | 0.2 | 5.9 | 0.1 | 9.3 | 0.1 | -4.1 | 0.1 | 61.8 | 0.5 | 31.3 | 0.9 | 30.6 | 1.0 |
| | | 87% | 16.3 | 0.1 | 4.0 | 0.1 | 9.6 | 0.1 | -6.2 | 0.1 | 79.5 | 0.4 | 41.4 | 0.5 | 38.1 | 0.7 |
| DLM - SIB | 40 | 0% | 9.6 | 0.1 | 9.8 | 0.2 | 9.0 | 0.1 | 0.8 | 0.2 | 0.1 | 0.3 | 6.6 | 1.7 | -6.5 | 1.7 |
| | | 22% | 11.8 | 0.1 | 8.9 | 0.2 | 8.8 | 0.1 | 0.1 | 0.3 | 24.5 | 1.5 | 25.2 | 1.2 | -0.7 | 2.1 |
| | | 46% | 15.1 | 0.2 | 8.3 | 0.2 | 8.3 | 0.1 | -0.1 | 0.2 | 45.0 | 1.1 | 44.6 | 0.7 | 0.4 | 1.4 |
| | | 68% | 21.0 | 0.1 | 6.7 | 0.2 | 7.2 | 0.1 | -0.4 | 0.2 | 68.0 | 1.0 | 65.9 | 0.4 | 2.1 | 1.1 |
| | | 87% | 33.0 | 0.2 | 4.2 | 0.2 | 5.3 | 0.1 | -1.1 | 0.2 | 87.3 | 0.6 | 84.0 | 0.3 | 3.3 | 0.6 |
| | 60 | 0% | 9.1 | 0.1 | 8.9 | 0.2 | 8.9 | 0.1 | 0.0 | 0.2 | 1.7 | 1.5 | 1.8 | 1.1 | 0.0 | 1.7 |
| | | 22% | 10.2 | 0.2 | 7.9 | 0.2 | 8.4 | 0.1 | -0.4 | 0.2 | 22.0 | 1.5 | 17.8 | 1.9 | 4.1 | 1.7 |
| | | 46% | 12.8 | 0.2 | 6.9 | 0.2 | 7.2 | 0.1 | -0.3 | 0.2 | 46.3 | 1.5 | 43.6 | 0.9 | 2.7 | 1.7 |
| | | 68% | 17.0 | 0.3 | 5.2 | 0.2 | 6.0 | 0.1 | -0.8 | 0.2 | 69.3 | 1.1 | 64.7 | 0.8 | 4.5 | 1.4 |
| | | 87% | 24.1 | 0.2 | 3.0 | 0.1 | 4.1 | 0.1 | -1.1 | 0.2 | 87.7 | 0.7 | 83.1 | 0.3 | 4.6 | 0.7 |
| | 80 | 0% | 9.6 | 0.1 | 9.7 | 0.3 | 9.3 | 0.1 | 0.5 | 0.3 | 0.7 | 1.5 | 3.0 | 1.6 | -2.3 | 2.3 |

| | | | | | | | | | | | | | | | | |
|---|---|---|---|---|---|---|---|---|---|---|---|---|---|---|---|---|
| | | 22% | 11.7 | 0.1 | 9.2 | 0.2 | 8.7 | 0.1 | 0.4 | 0.2 | 21.9 | 1.6 | 25.7 | 1.2 | -3.8 | 1.9 |
| | | 46% | 13.5 | 0.3 | 7.4 | 0.2 | 7.4 | 0.0 | 0.0 | 0.2 | 44.9 | 1.3 | 45.1 | 1.2 | -0.1 | 1.4 |
| | | 68% | 17.6 | 0.4 | 5.9 | 0.2 | 6.6 | 0.1 | -0.7 | 0.2 | 66.5 | 1.1 | 62.8 | 0.9 | 3.7 | 1.2 |
| | | 87% | 23.2 | 0.9 | 3.0 | 0.1 | 4.0 | 0.1 | -0.9 | 0.1 | 86.8 | 0.8 | 82.8 | 0.6 | 4.0 | 0.6 |
| CLM - TPR | 40 | 0% | 9.8 | 0.2 | 9.6 | 0.2 | 9.8 | 0.1 | -0.2 | 0.2 | 1.5 | 1.1 | 0.6 | 1.0 | 1.0 | 1.5 |
| | | 22% | 11.6 | 0.2 | 8.6 | 0.2 | 9.8 | 0.2 | -1.2 | 0.2 | 25.8 | 1.7 | 15.9 | 2.0 | 9.9 | 2.0 |
| | | 46% | 15.0 | 0.6 | 7.4 | 0.4 | 9.7 | 0.3 | -2.3 | 0.4 | 51.0 | 2.8 | 35.7 | 3.1 | 15.3 | 3.1 |
| | | 68% | 22.2 | 0.7 | 5.7 | 0.1 | 9.2 | 0.2 | -3.5 | 0.2 | 74.4 | 0.8 | 58.5 | 1.7 | 16 | 1.1 |
| | | 87% | 35.2 | 1.2 | 3.8 | 0.1 | 9.1 | 0.2 | -5.3 | 0.2 | 89.1 | 0.3 | 74.0 | 1.0 | 15.1 | 0.8 |
| | 60 | 0% | 9.5 | 0.1 | 9.3 | 0.2 | 9.2 | 0.1 | 0.1 | 0.2 | 2.2 | 1.3 | 3.2 | 2.0 | -1.0 | 2.1 |
| | | 22% | 10.4 | 0.2 | 8.4 | 0.1 | 9.3 | 0.2 | -0.9 | 0.2 | 19.6 | 1.4 | 10.7 | 2.2 | 8.9 | 1.9 |
| | | 46% | 14.6 | 0.5 | 6.9 | 0.2 | 9.0 | 0.1 | -2.1 | 0.2 | 52.5 | 1.6 | 38.3 | 2.1 | 14.3 | 1.5 |
| | | 68% | 20.2 | 0.7 | 5.7 | 0.1 | 9.1 | 0.2 | -3.4 | 0.2 | 71.6 | 1.1 | 54.9 | 2.0 | 16.7 | 1.4 |
| | | 87% | 29.2 | 1.2 | 3.6 | 0.1 | 8.9 | 0.1 | -5.3 | 0.1 | 87.7 | 0.5 | 69.4 | 1.4 | 18.2 | 1.0 |
| | 80 | 0% | 9.1 | 0.1 | 8.9 | 0.1 | 9.0 | 0.2 | -0.1 | 0.2 | 2.0 | 1.5 | 1.5 | 1.8 | 0.5 | 1.1 |
| | | 22% | 10.2 | 0.2 | 8.7 | 0.2 | 9.0 | 0.3 | -0.3 | 0.3 | 15.1 | 1.7 | 11.7 | 2.5 | 3.4 | 3.3 |
| | | 46% | 12.8 | 0.3 | 7.1 | 0.1 | 9.0 | 0.3 | -1.9 | 0.3 | 44.3 | 1.1 | 29.6 | 2.3 | 14.7 | 2.0 |
| | | 68% | 18.3 | 0.7 | 5.9 | 0.1 | 9.0 | 0.3 | -3.2 | 0.3 | 68.0 | 1.1 | 50.8 | 2.4 | 17.2 | 1.8 |
| | | 87% | 23.7 | 0.9 | 4.0 | 0.1 | 8.7 | 0.2 | -4.7 | 0.3 | 83.1 | 0.6 | 63.3 | 1.8 | 19.8 | 1.4 |
| DLM - TPR | 40 | 0% | 10.0 | 0.2 | 9.8 | 0.2 | 9.6 | 0.1 | 0.2 | 0.2 | 2.0 | 2.5 | 4.2 | 2.2 | -2.3 | 1.7 |
| | | 22% | 11.6 | 0.5 | 8.9 | 0.1 | 9.1 | 0.1 | -0.2 | 0.2 | 22.7 | 2.9 | 21.3 | 3.6 | 1.4 | 1.6 |
| | | 46% | 15.6 | 1.0 | 8.4 | 0.1 | 8.7 | 0.1 | -0.4 | 0.1 | 46.1 | 3.5 | 43.6 | 3.7 | 2.4 | 1.0 |
| | | 68% | 24.1 | 1.4 | 7.8 | 0.1 | 8.3 | 0.1 | -0.5 | 0.1 | 67.6 | 1.9 | 65.4 | 2.0 | 2.3 | 0.6 |
| | | 87% | 49.3 | 6.7 | 6.5 | 0.1 | 7.0 | 0.1 | -0.5 | 0.2 | 86.5 | 2.1 | 85.6 | 2.2 | 1.0 | 0.4 |
| | 60 | 0% | 9.3 | 0.1 | 9.4 | 0.1 | 9.4 | 0.2 | 0.0 | 0.2 | 0.0 | 0.2 | 0.1 | 0.3 | -0.1 | 0.4 |
| | | 22% | 11.3 | 0.5 | 8.9 | 0.1 | 9.2 | 0.2 | -0.3 | 0.2 | 21.2 | 3.9 | 18.5 | 3.8 | 2.7 | 2.0 |
| | | 46% | 15.0 | 0.9 | 8.3 | 0.1 | 8.8 | 0.2 | -0.5 | 0.2 | 44.5 | 3.2 | 41.1 | 3.6 | 3.5 | 1.2 |
| | | 68% | 21.2 | 1.6 | 7.3 | 0.1 | 8.4 | 0.1 | -1.1 | 0.1 | 65.3 | 2.9 | 60.3 | 3.1 | 5.0 | 0.7 |
| | | 87% | 49.3 | 3.3 | 5.9 | 0.1 | 6.5 | 0.1 | -0.7 | 0.2 | 88.1 | 0.7 | 86.7 | 0.9 | 1.4 | 0.4 |
| | 80 | 0% | 9.1 | 0.1 | 9.2 | 0.1 | 9.3 | 0.2 | -0.1 | 0.2 | 0.3 | 0.7 | 0.4 | 1.0 | -0.1 | 0.6 |
| | | 22% | 11.3 | 0.3 | 8.8 | 0.1 | 9.1 | 0.2 | -0.2 | 0.3 | 21.6 | 2.7 | 19.6 | 2.9 | 2.0 | 2.4 |
| | | 46% | 15.7 | 0.7 | 8.3 | 0.1 | 8.6 | 0.2 | -0.3 | 0.3 | 47.4 | 2.7 | 45.2 | 3.2 | 2.2 | 1.8 |
| | | 68% | 24.7 | 1.5 | 7.3 | 0.1 | 7.8 | 0.2 | -0.5 | 0.2 | 70.4 | 2.1 | 68.4 | 2.4 | 2.0 | 0.9 |
| | | 87% | 45.3 | 5.2 | 5.4 | 0.1 | 6.0 | 0.2 | -0.6 | 0.2 | 87.9 | 1.5 | 86.5 | 1.8 | 1.4 | 0.5 |
